# Beveled shaped axial oxygenator with improved hemodynamics

**DOI:** 10.1101/2024.03.19.24304532

**Authors:** B. Franke, L. Goubergrits

**Affiliations:** Institute of Computer-assisted Cardiovascular Medicine, Deutsches Herzzentrum der Charité, Berlin, Germany

**Keywords:** Oxygenator, flow design, manufacturing, hollow fiber membrane bundle, potting process

## Abstract

Oxygenators are a lifesaving technology used for blood oxygenation and decarboxylation in case of acute respiratory failure, chronic lung disease, and during open-heart surgery. Devices typically consist of a bundle of thousands of fiber membranes in a housing, with gas flowing inside the fibers and blood flowing in the opposite direction outside the fibers. Both ends of the fiber membranes are attached with an adhesive to prevent direct contact between gas and blood. The shape of the volume through which the blood flows is determined by the housing of the oxygenator and the internal end surfaces of the bonded parts of the fiber-membrane bundle. The traditional potting process results in a volume shape that is associated with stagnation zones, which are known to promote thrombus formation. In this study, an adapted potting process is proposed which results in a blood compartment with beveled end faces of the glued bundle parts. Using a numerical study, we have demonstrated that the novel oxygenator design results in optimized flow conditions.

## Introduction

Oxygenators or artificial membrane lungs are used for blood oxygenation and decarboxylation in case of acute respiratory failure, chronic lung disease, and during open heart surgery using heart-lung machines (cardiopulmonary bypass). The devices usually consist of hollow fiber-membrane systems in which blood flows around fibers and oxygen-mixed gas flows through the inner lumen of the fibers. Oxygenators use one of two main principles for blood and oxygen flow: Axial flow, where the blood flows parallel to the oxygen membranes, and crossflow, where the blood flows across to the oxygen membranes. In axial oxygenators, blood and oxygen flow in opposite directions, guaranteeing a steep oxygen gradient. Despite advances in material science, anticoagulation administration by coating, and optimization of the flow design, long-term use of oxygenators is limited due to lack of hemocompatibility [1]. Thrombus formation associated with regions of flow stagnation and/or recirculation is a major cause of device failure. Since the typical geometric design of oxygenators is a fiber-membranes bundle filled housing (e.g. tube) with an inlet and outlet for the blood located at the side, a non-optimal blood flow and of stagnation or low blood flow, and thus non-optimal blood oxygenation, are inevitable. This is a well-known challenge, and possible solutions have been proposed for some designs. For example, Hesselmann et al. proposed a biaxial centrifugation potting process generating an elliptical shape of the cured potting for the optimal flow in the stacked, cuboidal-shaped oxygenator Quadrox-i small adult (Getinge, Sweden) [2]. To solve this problem in a cylindrical housing with lateral blood inlet and outlet ports, we first analyzed the blood flow in an axial oxygenator using numerical simulations and then created an adapted design aimed at reducing the low-flow area/volume. Finally, we proposed a centrifugal manufacturing process to create the adapted shape of the inner form.

## Methods

Numerical simulations were used to analyse the blood flow through the cylindrical oxygenator. Due to the complex structure of the oxygenator, which consists of thousands of fiber-membranes, the design and simulation of a full-scale model is computationally expensive. However, the membranes form a repeating, somewhat homogeneous structure, and the influence of this structure on the blood flow can be simplified using a porosity model. Here the flow influencing properties of the porous model must be defined relative to the direction of flow, as axial flow and transverse flow are affected significantly differently by the fiber-membranes.

### CFD porosity model of the oxygenator flow

To identify the unknown parameters of the porosity model for both directions of flow, we created two miniaturized designs consisting of a few longitudinal or orthogonal fiber-membranes in a short cylinder (Fig. 1). The membranes had a diameter of 0.38 mm and were evenly distributed with a distance of 0.25 mm from one to another. The outer cylinder had a diameter of 3.4 mm and a length of 10 mm in the axial model and a diameter of 3.4 mm and a length of 4.0 mm in the transverse model. The fluid domains were extracted from the models and meshed in STAR CCM (v. 15.02.007, Siemens PLM Software Inc., Texas, USA) using a polyhedral mesh generator with a 3-cell boundary prism and a base size of 0.2 mm. This resulted in 508,021 cells for the longitudinal cylinder and 170,822 cells for the transverse cylinder (Fig. 1). The top and bottom surfaces of the cylinders were defined as the inlet and outlet boundaries, no-slip conditions were assumed on the outer surface and on the membrane surface. Blood was modeled as a Newtonian fluid with constant density of 1050 kg/m^3^ and a constant viscosity of 3.5 mPas. Turbulence was simulated by a Menter SST k-omega model, and a steady state approach was used.

**Figure 1:**
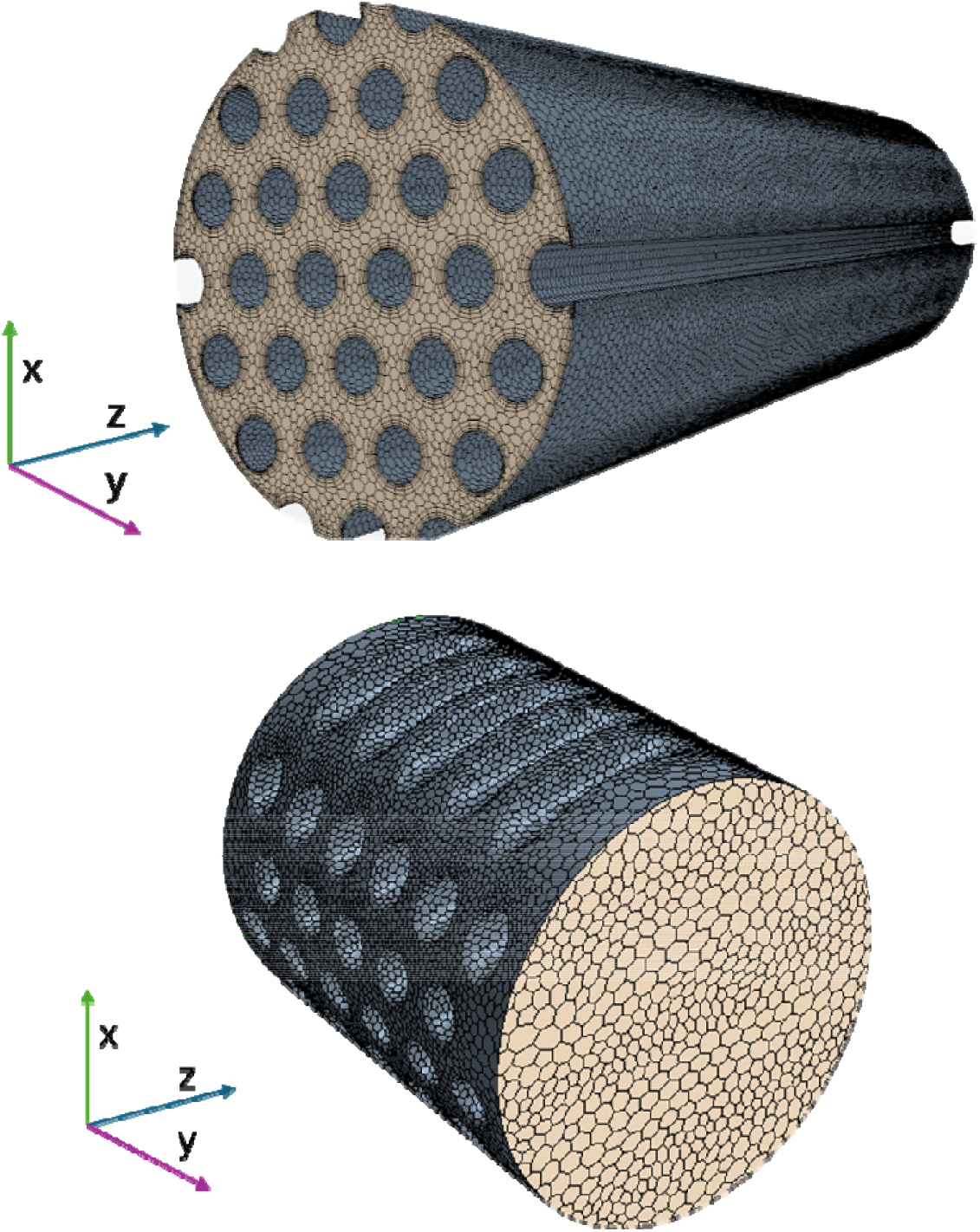
Volume mesh of the miniaturized designs representing axial (above) and transverse flow in an oxygenator.

For comparison, geometries of the longitudinal and transverse cylinders without fiber-membranes were generated and meshed according to the procedure described above. This resulted in 26,011 cells for the longitudinal cylinder and 9,072 cells for the transverse cylinder. The simulations of the cylindrical domains without fiber-membranes were done using a porosity model. The porosity was set to 0.6826 for the longitudinal cylinder and 0.7253 for the transverse cylinder, corresponding to the ratio between the fluid volumes of the fiber-membrane-filled and the unfilled cylinders. The resistance that the blood encounters when passing through the membranes was modeled by a porosity model based on Darcy’s Law, which describes the pressure loss (Δp) over a porous medium as a function of the velocity:

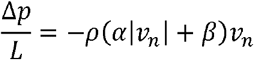

Here, ⍰_⍰_ is the artificial velocity that the fluid would have if it flowed through an empty cylinder, *α* is the porous inertial resistance, *β* is the porous viscous resistance and *ρ* is the density of the liquid.

In the solver used for the simulations, StarCCM+, this was implemented by using orthotropic tensors that define the internal resistance as P_i_ and the viscous resistance as P_v_. P_i_ and P_v_ consider the density of the fluid. We simulated blood flow in the fiber-membrane filled models and used the calculated Δp (calculated as the differences between the area averaged static pressure at the inlet and the outlet) to define P_i_ and P_v_. For this, flow rates of 0.1 ml/s, 1.0 ml/s, and 10.0 ml/s corresponding to an average cross-sectional velocity of 0.016 m/s, 0.16 m/s and 1.63 m/s in the longitudinal cylinder and 0.0176 m/s, 0.169 m/s and 1.72 m/s in the transverse cylinder were used. The validation of the porosity model pressure drop at flow rates of 0.3 ml/min and 3 ml/min was calculated. The results of these simulations, which show good applicability of the porous model, are summarized in Table 1. All simulations were performed in StarCCM+ software. The following values were determined for the porosity model:

**Table 1:**
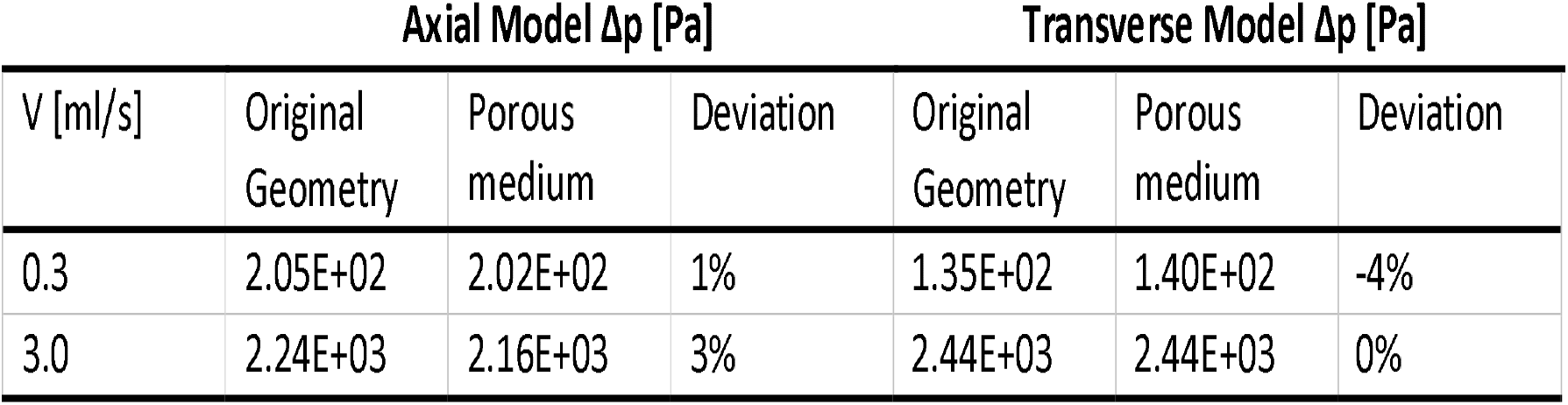
Δp in the original geometry and the porous medium of the axial and transverse model at different flow rates.

Porous Inertial Resistance P_i_ (X; Y; Z) = [2200000.0; 2200000.0; 50000.0] kg/m^4

Porous Viscous Resistance P_v_ (X; Y; Z) = [1000000.0; 1000000.0; 600000.0] kg/m^3 -s

The porosity model showed good accuracy compared to the original geometry in terms of Δp.

### CFD analysis of the oxygenator flow

After elaboration of the porosity model, the blood flow in a full-size oxygenator was simulated to compare two designs (classic vs. alternative). Both designs have the same cylindric outer shape as well as the same configuration of inlet and outlet ports for gas and blood (see figure 2). The basic oxygenator design (Figure 2, top) consists of a cylindrical shape with an inner diameter of 37 mm containing a bundle of approximately 4.860±200 fiber-membranes. The fiber-membrane bundle is fixed by an adhesive at both ends (Figure 2, blue colored parts). These glued parts of the bundle are also cylindrical in shape, forming a cylindrical compartment with a blood priming volume of 100 mm in length and 37 mm in diameter. Note, that the volume of the oxygenator compartment with blood is more than 1.000 times larger than the volume of the longitudinal cylinder used for the development of the porosity model, which required a mesh of 0.5 Mio cells. Respectively, an attempt to resolve the oxygenator with fiber-membranes would require a mesh of a huge size which is why the porosity approach was chosen.

**Figure 2:**
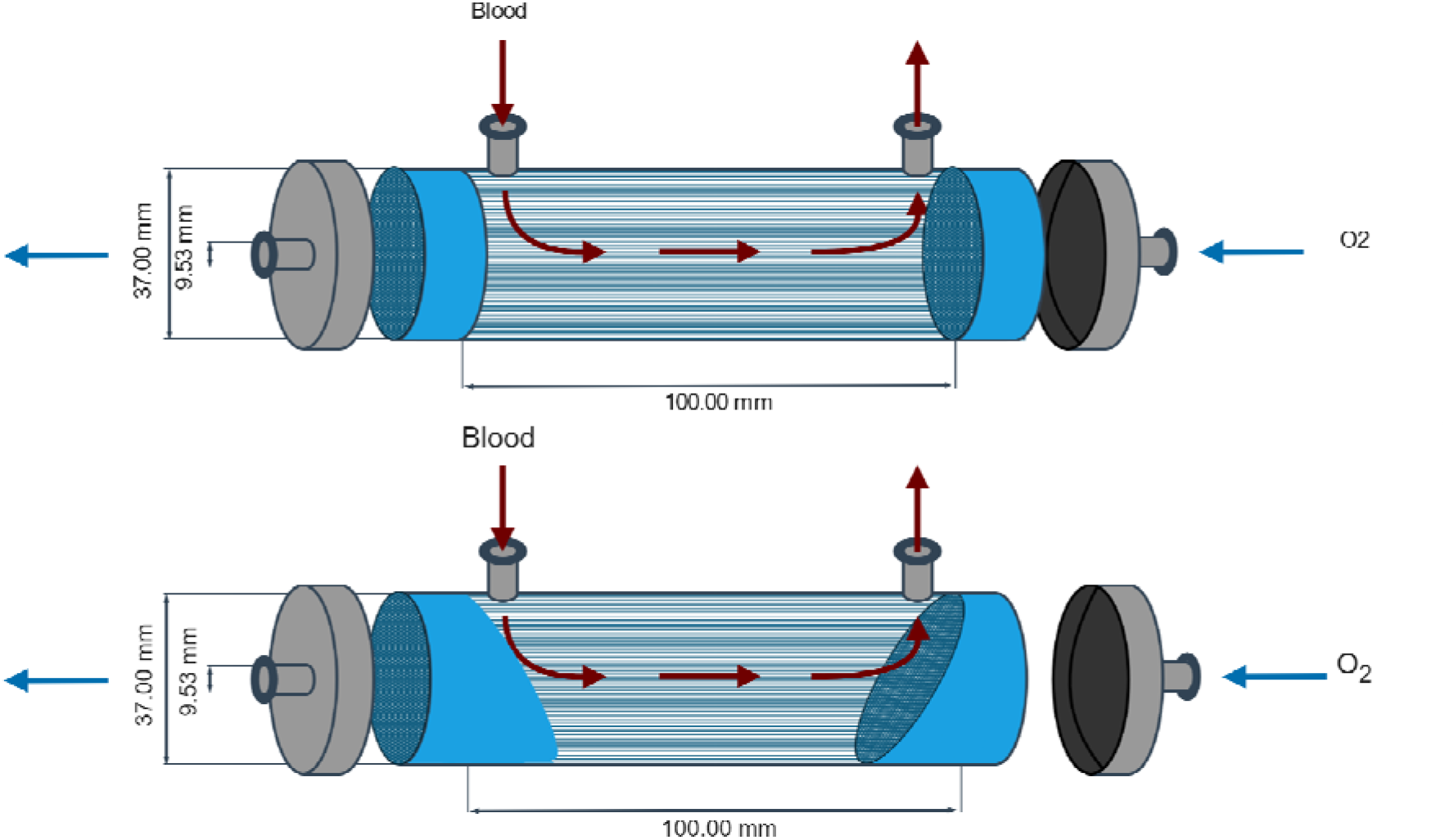
Top: Classic oxygenator model (cylindrically shaped housing). Bottom: Alternative novel design with beveled shaped potting.

In contrast, design 2 (Figure 2, bottom), which has the same number of fiber membranes as the classic design, uses a differently shaped adhesive part: The cylindrical parts have one end perpendicular to the axis of the oxygenator, while the opposite end, the inner end of the bonded part, is angled at 60°. The design is supposed to reduce areas of blood stagnation thus improving the performance of the oxygenator. Note that the outer design of the oxygenator including housing and ports remains unchanged. The average length of the blood path also remains the same at 100 mm. The blood flow inlet and outlet ports were 9.53 mm (3/8 inch) in diameter in both the Classic and Alternative models.

Note that the fabrication of the novel oxygenator device doesn’t require much adaptation. Traditionally in oxygenator manufacturing, membrane fibers are positioned longitudinally in the cylinder and the cylinder is filled with a potting compound (glue). The compound separates the gas and blood phases and seals the fiber ends. Since capillary forces would pull the compound up the fibers, a centrifugal force is typically used in the drying process to counteract the capillary forces [3]. To do this, the oxygenator housing cylinder is placed in a centrifuge so that the centrifugal force is perpendicular to the end-face of the cylinder (see figure 3), creating an angle of approximately 90° between the end-face and the side wall of the cylinder. However, for the alternative design consisting of a beveled glue-fixed part, the cylinder is placed offset 30° from the centrifugal normal. Thus, an axis of the cylinder is not parallel to the radial direction (Figure 3, red colored dashed lines).

**Figure 3:**
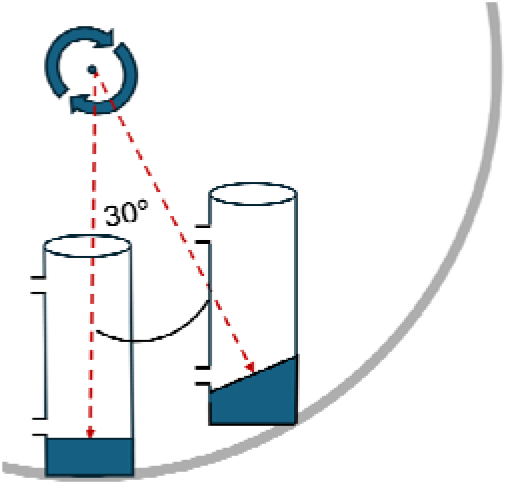
Potting process of classic model and beveled model using a centrifuge. In the beveled model, the cylinder is positioned 30° off the normal vector of the centrifuge.

To compare blood flow between the classic and the alternative novel model, the fluid volume was extracted from the geometrical models and meshed according to the above-described procedure, resulting in a cell number between 0.5 and 1.0 Mio. The fluid domain was defined as a porous region with a density of 0.6826 and a porosity model with the above parameters P_i_ and P_v_ was used to describe the resistance. The inlet flow rate was set to 100 ml/s, corresponding to a cardiac output of 6 l /min. A steady-state approach was chosen. A stopping criteria of 1000 iterative steps combine with a residual value less than 10^-5 for mass, momentum, turbulent kinetic energy and specific dissipation rate was set. To compare the flow fields in both models, first the vector field in the longitudinal section was qualitatively compared. Following, two cross sections were defined 20 mm proximal to the in- and outlet ports of the blood flow (see figure 4) and the distribution of velocity magnitudes was compared.

**Figure 4:**
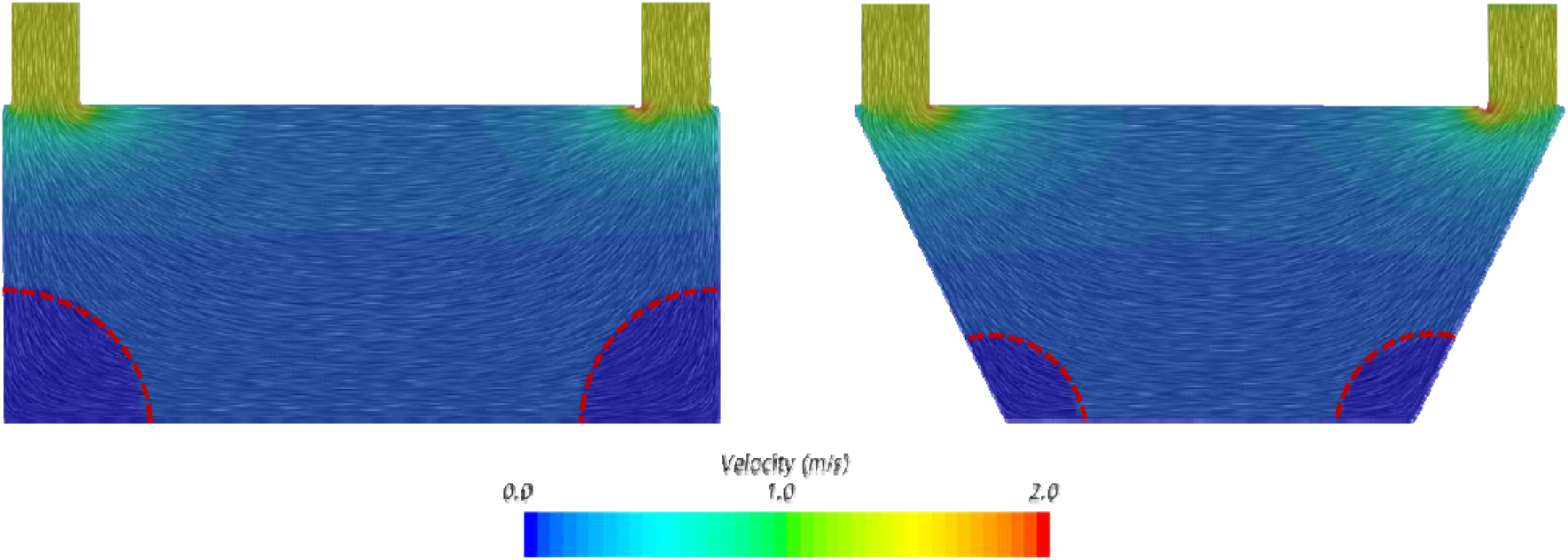
Vector field in the longitudinal section displayed as line integral convolution in the classic model (left) and the alternative model (right). Red lines mark areas with low flow conditions (velocity < 0.04 m/s).

## Results

Figure 4 shows the flow distribution in the longitudinal cross-section of the classic model (left) and in the novel model design (right). In both models, high velocities of up to 2 m/s were observed near the inlet and outlet ports. In the classic model, there were two regions opposite the inlet and outlet where the flow was very low (stagnant), indicated by velocities close to zero and marked as red dotted lines in figure 4. In the novel model, the lowest velocities also occurred near the wall opposite the inlet and outlet. However, the regions of low flow were much smaller than in the classic model (see figure 4). In the main region of the longitudinal section, the magnitude of the velocity was comparable between both models.

Next, quantitative analysis of the flow distribution was done by comparing the distribution of the velocity magnitudes in both models displayed as histograms in figure 5. In the classic model, mainly velocities between 0.0 and 0.3 m/s were calculated, with a dip in frequency at 0.05 m/s and a velocity of around 0.065 m/s being most frequent. In the novel model mainly velocities between 0.005 and 0.3 m/s were observed with velocity of around 0.05 m/s occurred most often. Overall, the frequencies of velocity magnitudes were more equally distributed in the novel model compared to the classic model.

**Figure 5:**
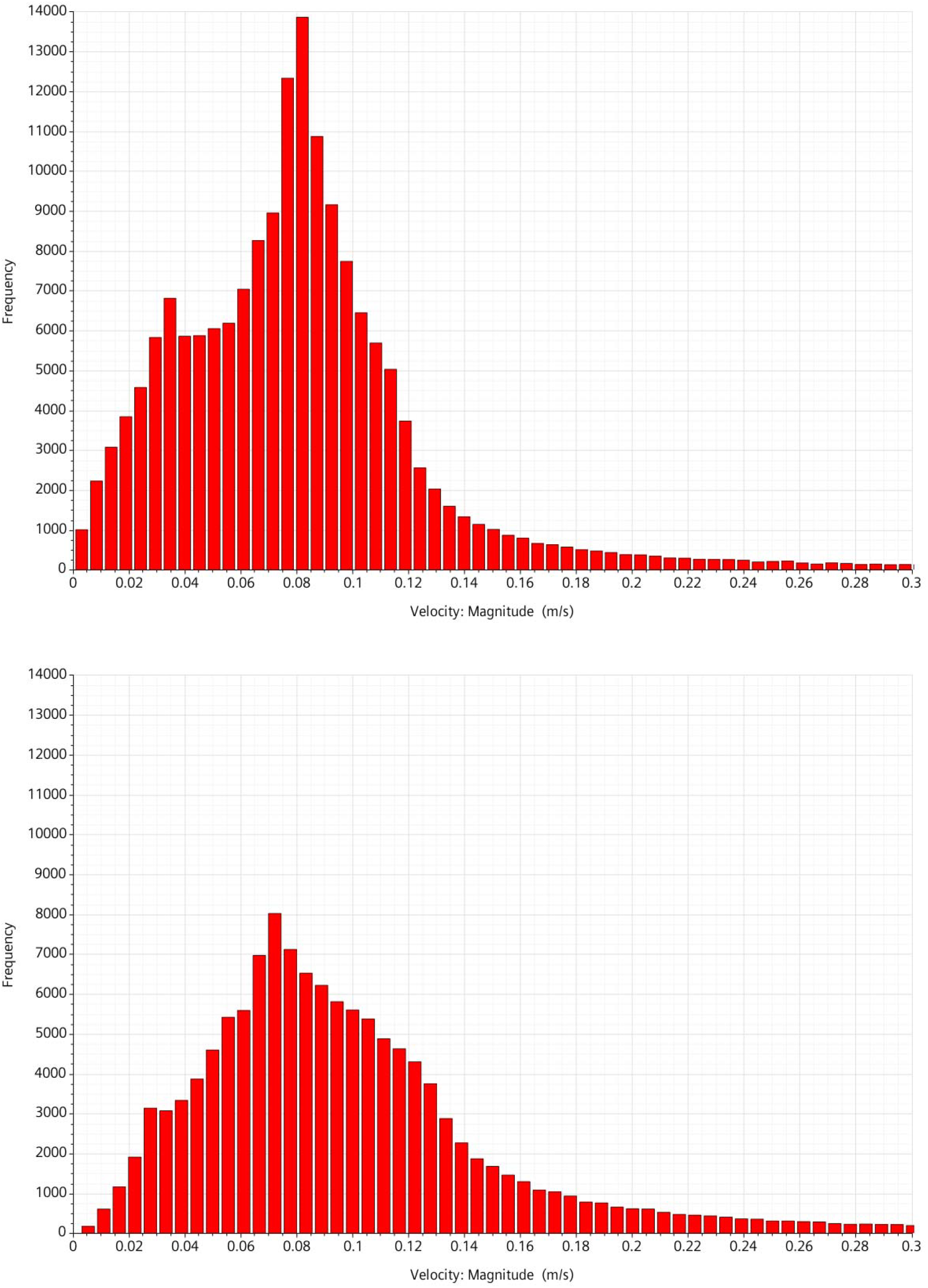
Distribution of velocity in the classic model (top) and the alternative model (bottom). Note that the upper limit of the velocity was set to 0.3 m/s for a better overview to although higher velocities up to 2 m/s exist at the inlet and outlet.

## Discussion

Optimal flow conditions in oxygenators which avoid non-physiologic conditions and thus thrombus formation are important to improve long-term performance and increase safety and efficacy of devices. Thrombosis in oxygenators is still one of the major problems limiting broader and long-term use of these artificial organs [4-7]. Further, optimal flow conditions improve oxygen saturation and thus enable size reduction. However, design optimization cannot be done without considering the reliability of manufacturing processes and, specifically in our case, the potting process. The novel design combined with a slightly modified potting process fulfills all these requirements. The proposed potting process was not tested experimentally. However, an earlier experimental study by Hesselmann et al [2] showed that it is feasible to produce adhesive-bonded parts with complex shapes. The beveled shape of the glued fiber membrane bundle reduces regions with stagnant flow thus resulting in a more evenly distributed flow field. Furthermore, the proposed design also reduced the blood priming volume, which is known to be associated with another problem of oxygenators use – acute kidney injury [8, 9]. Due to optimal flow conditions, we are expecting an improved oxygen transfer with a shorter saturation length. This should be proved, however, in future studies.

The current numerical study is limited to (1) modeling blood as a single-phase fluid with constant density and viscosity without modeling gas or oxygen saturation, and (2) using a homogeneous porosity model for the entire blood domain without resolving the fibrous membranes, which undoubtedly affect the flow field inhomogeneously. The porosity model used in this study was verified regarding the calculated pressure drop. Furthermore, a set of earlier studies showed the usability of the porosity model for studying blood flow in oxygenators [10, 11]. The study presented is a preliminary feasibility study that demonstrates the general possibility of flow optimization through the beveled shape of the bonded parts. Other potential optimizations could be related to the position of the blood ports or the angle of the beveled surface.

## Conclusion

A novel design of a classic cylindrically shaped oxygenator with a modified potting process for fixation of the fiber membranes bundle was proposed. The numerical proof-of-concept study confirmed the optimization of the blood flow field which results in a reduction of regions with blood stagnation in the corners of the housing opposite to the inlet and outlet ports, which are associated with a higher risk of thrombus formation. The novel design may emerge novel oxygenator devices with an improved performance, which could result in better long-term stability and efficiency.

## Data Availability

All data produced in the present study are available upon reasonable request to the authors

## Declarations

### Ethical approval

This article does not contain any studies with human or animal subjects performed by any of the authors.

